# Suboptimal dietary patterns are associated with accelerated biological aging in young adulthood: a twin study

**DOI:** 10.1101/2024.06.25.24309391

**Authors:** Suvi Ravi, Anna Kankaanpää, Leonie H. Bogl, Aino Heikkinen, Kirsi H. Pietiläinen, Jaakko Kaprio, Miina Ollikainen, Elina Sillanpää

**Author notes:** Corresponding author (S.R.). (A.K.); (E.S.); (L.H.B.); (A.H.); (J.K.); (M.O.).

## Abstract

**Background & aims:** Suboptimal diets increase morbidity and mortality risk. Epigenetic clocks are algorithms that can assess health and lifespan, even at a young age, before clinical manifestations of diseases. We investigated the association between dietary patterns and biological aging in young adult twins.

**Methods:** The data were drawn from the population-based FinnTwin12 study and consisted of twins aged 21–25 years (n=826). Food and beverage intakes were assessed using a food frequency questionnaire. Biological aging was estimated using the epigenetic clocks GrimAge and DunedinPACE. Latent class analysis was used to identify dietary patterns. The association between dietary patterns and biological aging was assessed using linear regression modeling at the individual level, followed by within–twin pair analyses to account for genetic liabilities and shared familial confounders.

**Results:** Six dietary patterns were identified: 1) High fast food, low fruits and vegetables (F&V), 2) Plant-based, 3) Health-conscious, 4) Western with infrequent fish, 5) Western with regular fish, and 6) Balanced average. At the individual level, GrimAge acceleration was slower in the Plant-based, Health-conscious, and Balanced-average patterns compared to the High fast food, low F&V, and faster in the Western with infrequent fish pattern compared to the Balanced average, regardless of sex, nonalcoholic energy intake, smoking, and alcohol consumption. After further adjustment for BMI and sports participation, the strengths of the associations modestly decreased; however, the difference between the Balanced-average and High fast food, low F&V patterns remained significant. The pace of aging (DunedinPACE) was slower in the Plant-based pattern compared to the High fast food, low F&V and the Western with infrequent fish patterns after adjustment for sex, nonalcoholic energy intake, smoking, and alcohol. The effect sizes were attenuated and reached a non-significant level when BMI and sports participation were added to the model. Most of the associations were replicated in the within-pair analyses among all twin pairs and among dizygotic twin pairs, but the effect sizes tended to be smaller among monozygotic twin pairs. This suggests that genetics, but not a shared environment, may partially explain the observed associations between diet and biological aging.

**Conclusion:** Diets high in fast food, processed red meat, and sugar-sweetened beverages and low in fruits and vegetables are associated with accelerated biological aging in young adulthood. The clustering effect of lifestyle factors and genetic confounders should be considered when interpreting the findings.

## INTRODUCTION

Diet quality is a modifiable lifestyle factor that significantly impacts mortality and morbidity risks (1–3). Diet-related noncommunicable diseases, such as cardiovascular diseases and type 2 diabetes, are among the leading causes of death worldwide (4,5). Health-promoting diets—that is, those rich in vegetables, fruits, legumes, nuts, whole grains, vegetable oils, and fish and low in red and processed meat, high-fat dairy, and refined carbohydrates—play a crucial role in reducing the risk of these diseases (2,3) and all-cause mortality (1).

One mechanism by which diet can influence health and lifespan involves changes in the epigenome, which further modify pathways leading to diseases (6). Epigenetic modifications alter gene expression without changing the DNA sequence (7). The most well-established and valuable epigenetic marker in human disease studies is DNA methylation (DNAm) (8,9). This marker typically involves the attachment of a methyl group to a cytosine-phosphate-guanine (CpG) dinucleotide on a DNA strand (10). The activity of DNAm varies depending on its genomic location. Methylation of DNA promoters often suppresses gene expression, whereas methylation of gene bodies can enhance gene expression (11–13). Although genetic factors contribute to DNAm, environmental and lifestyle factors also play a significant role in this process (14). Genetic factors seem to play a larger role in young individuals, whereas environmental and lifestyle factors explain more of the variation in DNAm in older cohorts (14).

Epigenetic alterations are one of the classical hallmarks of aging (15), and DNAm status at a set of specific CpG sites can be used to estimate individuals’ biological age. To date, several epigenetic clocks (i.e., machine learning algorithms that utilize different CpG sites) have been validated to predict aging-related phenotypes (16,17). The first two widely used clocks, the Hannum and Horvath clocks, were developed to estimate chronological age (18,19). More recent “second- and third-generation clocks,” GrimAge and DunedinPACE, were developed to predict lifespan and the pace of aging, respectively, rather than chronological age (20,21). For GrimAge, the difference between the estimated biological age and chronological age indicates age acceleration, whereas DunedinPACE directly provides an index of the pace of aging—that is, an average rate of years of biological aging per year of chronological aging. Increased age acceleration and an increased pace of aging have been shown to predict morbidity and mortality in several cohorts (20–24).

Some observational studies have found a relationship between higher diet quality and age deceleration (25–29). However, most current evidence comes from middle-aged and older adults. During young adulthood (between the ages of 18 and 35), individuals transition from adolescence to a stage where they begin to manage the responsibilities associated with independent living (30). This phase of life may involve leaving the parental home, starting work or academic education, and beginning a relationship or parenthood, all of which have the potential to affect dietary patterns (30–32). It has been reported that diet quality tends to decrease during the transition from adolescence to young adulthood (33). As diseases usually take decades to manifest, it is important for disease prevention to investigate the link between diet and health from an early age before clinical signs of age-related diseases appear. Epigenetic clocks, especially newer ones, emerge as a potential metric for this purpose.

Previous studies examining the relationship between diet and biological aging have not accounted for genetic liability or shared familial confounders. In addition, previous studies investigating the association between diet and biological aging have used theory-based *a priori* methods, i.e., dietary indices, to asses diet quality (25–28). Instead, *a posteriori* methods provide a hypothesis-free, empirically informed approach to analyzing dietary patterns, which are the overall combinations of foods and beverages typically consumed (3). Latent class analysis (LCA) is one such person-oriented, data-driven method used to identify unobserved subgroups within a population based on patterns of responses across multiple observed variables (34). In this paper, we investigated the association between LCA-derived dietary patterns and biological aging in young adult twins. In addition to individual-level analyses, we employed within–twin pair analyses to account for shared environment and genetics.

## MATERIALS AND METHODS

### Study design and participants

This study utilized data from the fourth wave of the intensively studied sample of the FinnTwin12 study, collected between 2006 and 2009. FinnTwin12 is a population-based longitudinal study comprising Finnish twins born between 1983 and 1987 (35,36). The intensively studied sample (1035 families) was created through random sampling from each birth cohort when the twins were 11–12 years old. The sample was enriched to include individuals at familial risk of alcoholism. Specifically, all remaining twins in each cohort, where one or both parents exceeded the threshold on a questionnaire assessing alcoholism (the Malmö-modified Michigan Alcoholism Screening Test; Mm-MAST 37), were added to the randomly selected sample. These families comprised 28% of the overall sample. At the age of 14, 1852 twins were interviewed and later invited to participate in the fourth study wave as young adults.

The data for the fourth wave were gathered during in-person study visits when the twins were between the ages of 20 and 26 years. A total of 1347 twins (73% participation rate) underwent assessments at this phase. Dietary data were available for 1294 participants. Of these, 839 also had epigenome data from blood samples. Four participants later withdrew their consent to participate in the study, eight participants were excluded due to incomplete dietary data (>8 items missing in the food frequency questionnaire), and one was excluded due to implausibly low energy intake (<600 kcal/day in females). This resulted in a final sample size of 826 for the individual analyses. For the within–twin pair analyses, 100 individuals for whom we did not have valid data on their cotwin were excluded from the data, resulting in a sample of 726 individuals—that is, 363 complete twin pairs of which 206 were dizygotic (DZ) and 157 monozygotic (MZ).

All participants provided written informed consent. The data were collected according to the principles of the Declaration of Helsinki, and the study was approved by the Institutional Review Board of Indiana University and the ethics committees of the University of Helsinki and Helsinki University Central Hospital (113/E3/2001 and 346/E0/05).

### Dietary intake

Habitual dietary intake was assessed through a food frequency questionnaire containing 52 foods and nonalcoholic beverages. A detailed description of the dietary assessments was provided elsewhere (38). Briefly, the participants were asked about the typical frequency of 52 foods and nonalcoholic beverages consumed during the previous 12 months. Additionally, they reported the number of slices of rye bread, whole grain bread, and white bread consumed daily. The participants also reported the amount of fat spread per slice of bread. The intake of each food and beverage item in grams per day and the daily macronutrient and nonalcoholic energy intake were calculated using the national computer-based food composition database Fineli (www.fineli.fi).

### DNA methylation and biological age

Genomic DNA was isolated from venous blood samples using commercial kits. DNA samples of 1 µg were then bisulfite-converted using an EZ-96 DNA Methylation-Gold Kit (Zymo Research, Irvine, CA) following the manufacturer’s instructions. The samples from both twins were distributed on the same plate to minimize potential batch effects. Measurements of DNA methylation were performed using Illumina’s Infinium HumanMethylation450 BeadChip (covering over 450,000 CpGs) for 742 samples and the Infinium MethylationEPIC BeadChip (covering 850,000 CpGs; Illumina, San Diego, CA) for 84 samples. Methylation data were combined and preprocessed using the R package *meffil* (39). The preprocessing steps have been previously described in detail (40).

Two epigenetic clocks—namely, principal component (PC)–based GrimAge (20,41) and DunedinPACE (21)—were used in this study. GrimAge, established in 2019, uses 1030 CpGs to predict time-to-death and is a composite measure incorporating chronological age, sex, and DNAm-based surrogate biomarkers related to seven plasma proteins and smoking pack-years (20). To increase reproducibility, we utilized the PC version of GrimAge (PC-GrimAge) in the analysis because it is less sensitive to technical noise arising from DNAm data compared to the original DNAm GrimAge (41). DunedinPACE, established in 2022, is an updated version of the 2020 clock DunedinPoAm (42). It was trained to predict changes in 19 biomarkers from age 26 to age 45, utilizing 173 CpGs to forecast the pace of aging (21).

PC-GrimAge and DunedinPACE estimates were produced using R packages (https://github.com/MorganLevineLab/PC-Clocks and https://github.com/danbelsky/DunedinPACE, respectively; 18,19). PC-GrimAge acceleration (AAPC-Grim), which reflects the difference between chronological age and biological age assessed by PC-GrimAge, was calculated as the residual from a regression model in which the estimated biological age was regressed on chronological age. The epigenetic aging measures were screened for outliers (values more than 5 standard deviations from the mean). No outliers were detected.

### Lifestyle-related covariates

BMI (kg/m^2^) was calculated using the height and weight measured at the study visit. Sports-related physical activity was assessed using the Baecke questionnaire (43). The participants reported the number of months per year and hours per week they participated in their two most regularly practiced types of exercise. The sport index was calculated based on the volumes and intensities of the reported activities, as described by Baecke et al. (43) and Mustelin et al. (44).

Smoking status, categorized as never smokers, former smokers, occasional smokers, and daily smokers, was self-reported by the participants. The number of servings of beer, wine, spirits, and other alcoholic beverages consumed during a typical week was assessed through a detailed interview as part of the Semi-Structured Assessment of Genetics of Alcoholism (SSAGA; 45). To calculate alcohol intake in grams, the number of servings of each beverage type consumed per week was multiplied by the respective portion sizes and alcohol content. This yielded the total weekly alcohol intake. Subsequently, the weekly alcohol intake in grams was divided by 7 to determine the average daily intake.

### Statistical analyses

We employed LCA to identify different dietary patterns. The intake of each food and nonalcoholic beverage item was used as an indicator variable. Prior to conducting LCA, the items were log-transformed due to their skewed distributions. For indicators with a strong floor effect (8 out of 55 indicators), the variables were recoded as binary, indicating whether or not the participants consumed the corresponding food item. An LCA model with 1–7 classes was fitted. The classification was based on the means and variations of the continuous indicator variables and the conditional item probabilities of the binary indicators. In each step, we used the following indices to evaluate the goodness of fit: Akaike’s information criterion, the Bayesian information criterion (BIC), and the sample size–adjusted BIC (aBIC). Lower values of the information criteria suggest a better fit for the model. Moreover, we used likelihood-based tests—namely, the Vuong–Lo– Mendell–Rubin likelihood ratio test (VLMR-LRT) and the Lo–Mendell–Rubin (LMR) test—to assess whether adding a class led to a statistically significant improvement in model fit. A low p-value suggests that the model with one class less should be rejected. In each step, classification quality was assessed using the average posterior probabilities for the likeliest latent class membership (AvePP) and the entropy values. Entropy values higher than 0.8 indicate that the classification uncertainty is low (46). AvePPs close to 1 indicate a clear classification. To simplify further analysis, the classification error was ignored, and each participant was assigned to the class for which the posterior membership probability was the largest (modal assignment).

Linear regression modeling was conducted to study the differences in biological aging between classes with different dietary patterns. Covariates were sequentially added to the model. The standard errors were corrected for nested sampling within families using the “type=complex” option in Mplus. Following these individual-level analyses, the associations were estimated at the within– twin pair level using multilevel models in all twin pairs and separately for MZ and DZ pairs (47). This approach addresses shared environmental factors, either measured or unmeasured, across all twin pairs. Additionally, within MZ pairs, which share all their genetic variants, this method accounts for genetic influences, while in DZ twin comparisons, it addresses, on average, 50% of the segregating genetic influences.

We also assessed whether sex, nonalcoholic energy intake, smoking status, alcohol consumption, BMI, or sports participation may modify the impact of diet on biological aging by incorporating an interaction term into our models. The Wald test was used to test the significance of the interaction term.

Descriptive statistics were analyzed using SPSS version 28.0 (Armonk, NY: IBM Corp). LCA and regression modeling were conducted using Mplus software version 8.2 (48). The parameters of the models were estimated using the full information maximum likelihood (FIML) method with robust standard errors. This approach produces unbiased parameter estimates when data are missing at random (MAR). In all statistical tests, the alpha level of significance was set at 0.05.

## RESULTS

The demographic and dietary characteristics of the whole sample and stratified by sex are presented in Table 1. On average, the participants were 22.4 (0.7) years old, with an age range of 21–25 years, with 58.2% being female. Compared to females, males had a higher BMI and alcohol intake and smoked more frequently. In addition, males exhibited higher absolute energy and macronutrient intakes, while females consumed a higher percentage of carbohydrates relative to their total energy intakes.

**Table 1.**
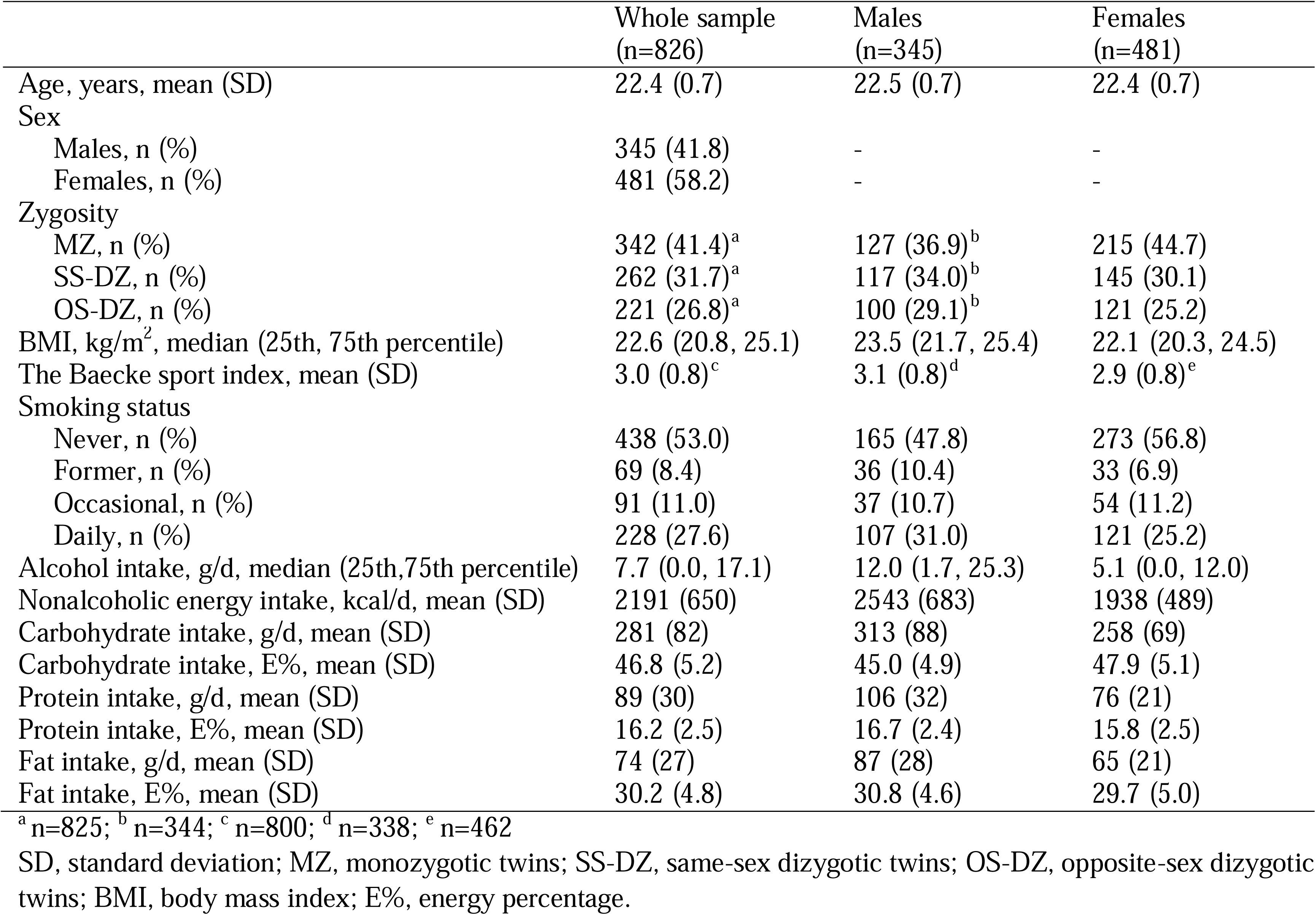
Demographic and dietary characteristics of the whole sample and stratified by sex.

The LCA models with 1 to 7 classes are shown in Table S1. The VLMR-LRT and LMR test indicated that even a 2-class model would be sufficient. However, the model fit improved based on the information criteria until the inclusion of the sixth class in the model. The entropy reached its maximum level in the sixth step as well. Consequently, a 6-class solution was selected for further analysis.

Fig. 1 shows the mean consumption of each food for different dietary patterns. The probabilities of consumption of different food items treated as binary variables in the LCA are presented in a separate figure in the Supplement (Fig. S1). We named the dietary patterns as follows: 1) High fast food, low fruits and vegetables (F&V), 2) Plant-based, 3) Health-conscious, 4) Western with infrequent fish, 5) Western with regular fish, and 6) Balanced average.

**Fig. 1.**
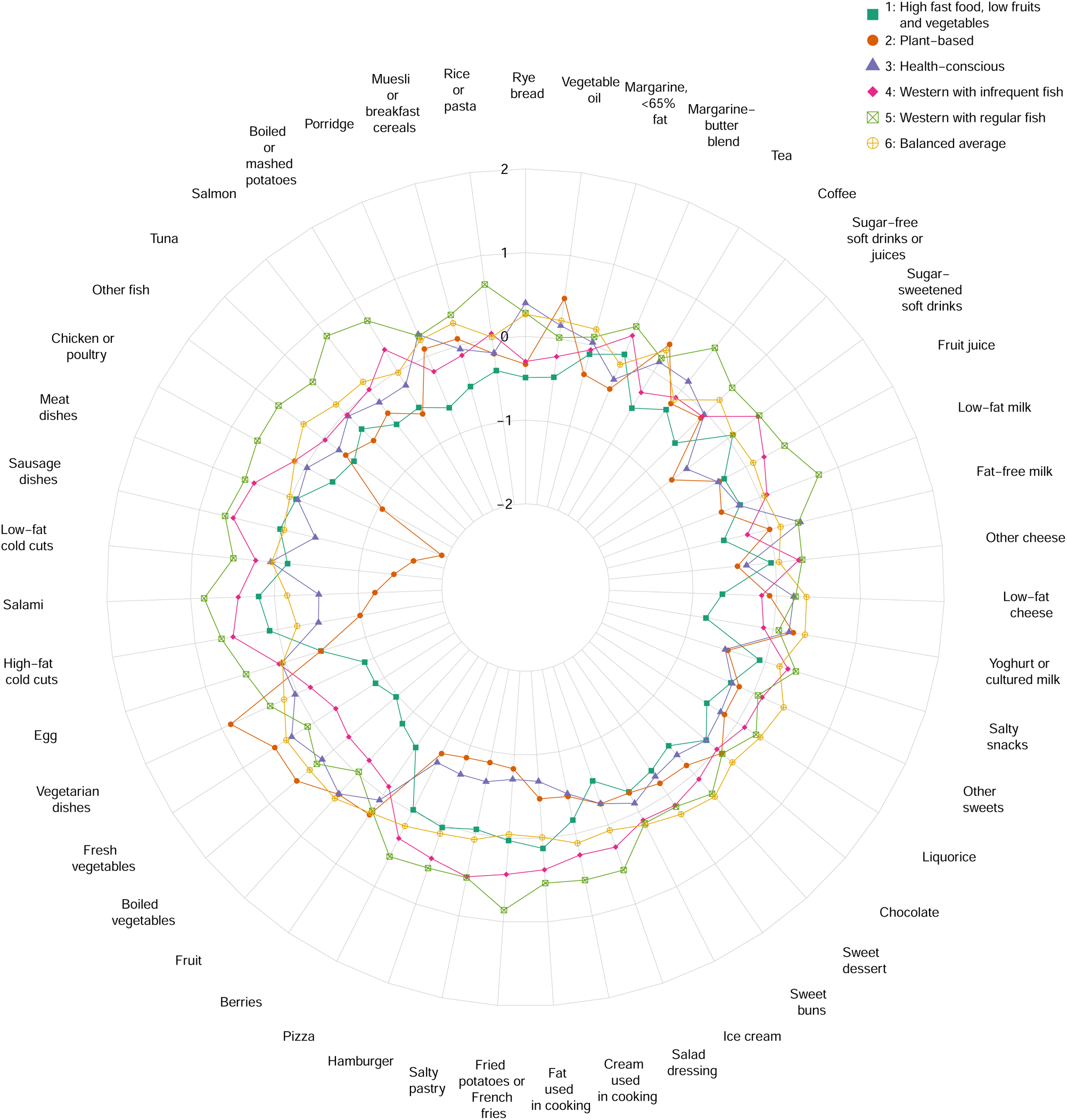
Mean consumption of selected food items by dietary patterns derived from latent class analysis. The mean values are presented on a Z-score scale.

The High fast food, low F&V pattern was characterized by relatively high intakes of high-fat, processed meat, fast food (i.e., hamburgers, pizza, pastry, fried potatoes, and French fries), sugar-sweetened beverages, and margarine-butter blend. In turn, individuals with this dietary pattern consumed lower quantities of vegetables, fruits, berries, grain products, fish, yogurt, low-fat cheese, fat-free milk, and vegetable oils. Overall, participants in the High fast food, low F&V pattern reported a relatively low frequency of food item consumption.

The participants in the Plant-based pattern tended to consume high amounts of vegetables, fruits, berries, porridge, muesli or cereals, chocolate, yogurt, tea, and vegetable oils and low quantities of fast food, fish, meat, boiled or mashed potatoes, and soft drinks.

The participants in the Health-conscious pattern favored low-fat products, such as low-fat meat, fat-free milk, and low-fat cheese. Their intake of vegetables, fruits, berries, rye bread, and porridge was also high. In turn, the intake of fast food, high-fat meat, soft drinks, and fruit juices was low in the Health-conscious pattern.

The Western with infrequent fish pattern was characterized by high intakes of meat, fast food, soft drinks, and margarine-butter blend and lower intakes of rye bread, porridge, boiled potatoes, vegetables, fruits, berries, yogurt, low-fat cheese, fish, and vegetable oils. This pattern was relatively similar to the High fast food, low F&V pattern; however, the latter consumed even lower amounts of vegetables, fruits, and berries.

The participants with a Western in regular fish dietary pattern consumed high amounts of rice or pasta, meat, fish, fast food, sugar-free drinks, soft drinks, fruit juice, low-fat milk, coffee, and margarine-butter blend and relatively low amounts of vegetables, fruits, berries, desserts, and vegetable oils. Overall, the participants in this pattern reported high intakes of different food items. Thus, the absolute and relative intakes of vegetables, fruits, and berries were higher in this pattern than in the High fast food, low F&V pattern.

Finally, the participants in the Balanced-average dietary pattern tended to consume high amounts of rye bread, porridge, fish, vegetables, fruits, berries, desserts, and tea and lower amounts of high-fat meat, margarine-butter blend, and coffee, but the intake of other food and beverage items was average.

The characteristics of the participants with different dietary patterns are presented in Table S2. In general, males tended to be in the majority in the Western with infrequent fish and Western with regular fish dietary patterns; these groups were also characterized by the highest BMIs, alcohol consumption, and nonalcoholic energy and fat intakes. Instead, females predominated in the groups with Plant-based, Health-conscious, and Balanced-average dietary patterns. These patterns included the highest rates of never smokers and were characterized by relatively low alcohol intake and higher carbohydrate consumption relative to the total energy intake.

The individual-level associations between dietary patterns and biological aging among the entire sample are presented in Fig. 2. The High fast food, low F&V pattern was treated as the reference group, as it exhibited the highest age acceleration and pace of aging in Model 1 (adjusted for family relatedness, daily nonalcoholic energy intake, and sex). Compared to the High fast food, low F&V pattern, AAPC-Grim was slower in the Plant-based (β=-0.122, 95% confidence interval [CI] −0.207, −0.037), Health-conscious (β=-0.171, 95% CI −0.269, −0.073), and Balanced-average patterns (β=-0.198, 95% CI −0.298, −0.099) in Model 1. After further adjustment for smoking status and alcohol intake (Model 2), the strength of these associations decreased but remained statistically significant (β=-0.077, 95% CI −0.152, −0.002; β=-0.112, 95% CI −0.200, −0.025; and β=-0.148, 95% CI −0.236, −0.061, respectively). After additional adjustment for the BMI and the Baecke sport index (Model 3), the strength of the associations was further attenuated, and the associations of the Plant-based and Balanced-average patterns with the High fast food, low F&V pattern became statistically insignificant (β=-0.059, 95% CI −0.133, 0.016; β=-0.087, 95% CI −0.178, 0.005, respectively). However, the association between the Balanced-average and the High fast food, low F&V patterns remained statistically significant in Model 3 (β=-0.131, 95% CI −0.218, −0.044; Fig. 2).

**Fig. 2.**
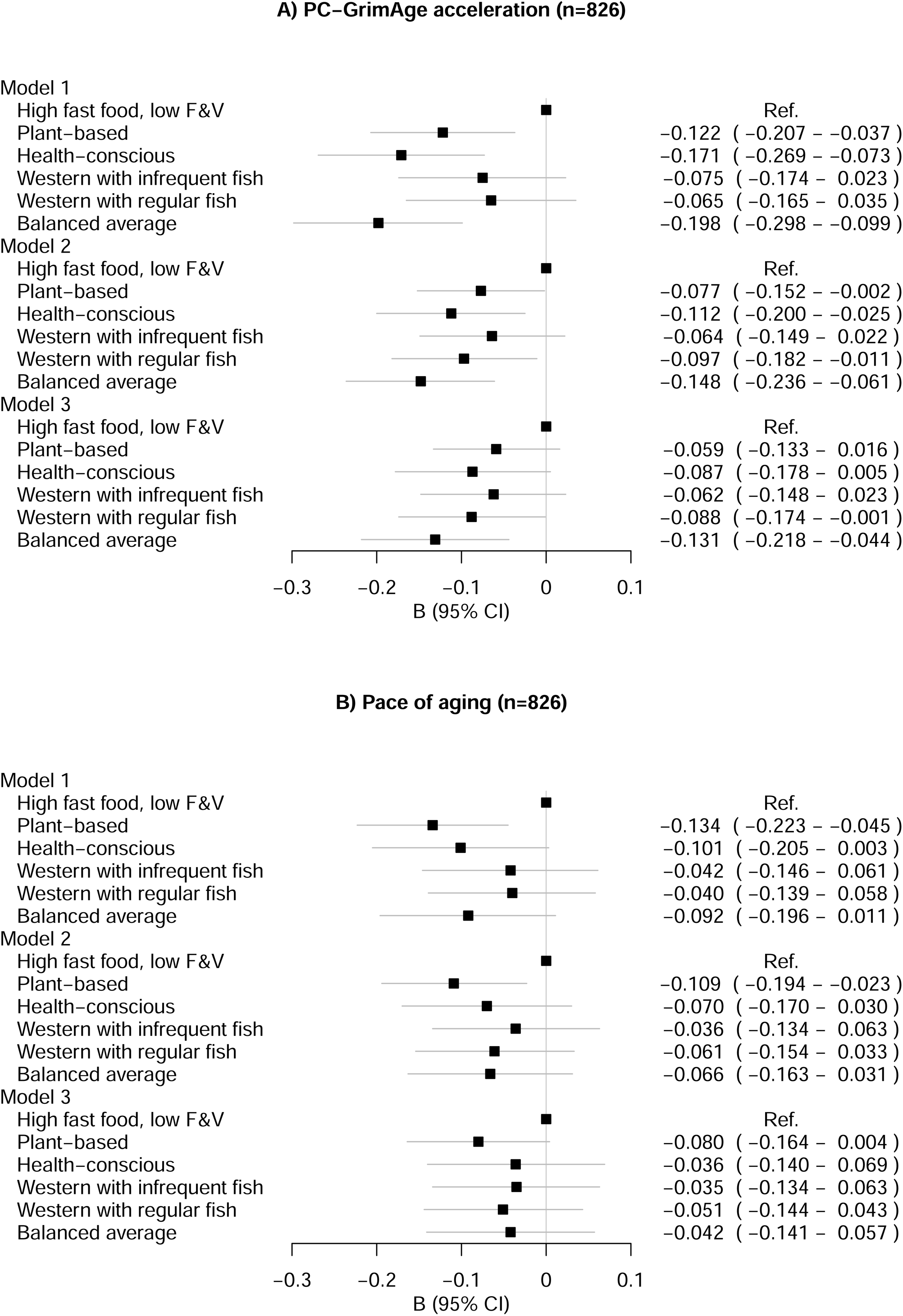
Differences between the dietary patterns in A) principal component-based GrimAge acceleration and B) the pace of aging (measured by the DunedinPACE estimator): individual-level analysis. Model 1 was adjusted for family relatedness, total nonalcoholic energy intake, and sex; Model 2 was adjusted for the covariates in Model 1 plus smoking status and daily alcohol intake; and Model 3 was adjusted for the covariates in Model 2 plus BMI and the Baecke sport index. F&V, fruits and vegetables; B, standardized regression coefficient; CI, confidence interval; Ref., reference group.

In addition to using the High fast food, low F&V pattern as a reference group, we also conducted pairwise comparisons with other classes serving as the reference. In these comparisons, AAPC-Grim was faster in the Western with infrequent fish pattern compared to the Balanced-average pattern in Models 1 and 2 (β=0.134, 95% CI 0.039, 0.229 and β=0.093, 95% CI 0.004, 0.183, respectively; results not shown). In Model 3, the effect size continued to attenuate from Models 1 and 2, with the association reaching a non-significant level (β=0.077, 95% CI −0.010, 0.164). In addition, in Model 1, AAPC-Grim was slower in the Health-conscious pattern compared to the group with a Western with infrequent fish dietary pattern (β=-0.100, 95% CI −0.188, −0.011). The effect sizes were attenuated in Models 2 and 3, with the association reaching a non-significant level (β=-0.052, 95% CI −0.139, 0.035 and β=-0.028, 95% CI −0.116, 0.061 in Models 2 and 3, respectively). Furthermore, AAPC-Grim was faster in the group with a Western with regular fish pattern than in that with a Balanced-average pattern in Model 1 (β=0.104, 95% CI 0.011, 0.197), with the effect sizes reducing to statistically insignificant levels in Models 2 (β=0.030, 95% CI - 0.056, 0.115) and 3 (β=0.024, 95% CI −0.059, 0.107). No other statistically significant differences were observed in pairwise comparisons for AAPC-Grim.

Regarding DunedinPACE, the only significant difference from the High fast food, low F&V pattern was observed in the Plant-based pattern, which exhibited a slower pace of aging in Models 1 and 2 (β=-0.134, 95% CI −0.223, −0.045 and β=-0.109, 95% CI −0.194, −0.023, respectively). The strength of this association was modestly attenuated and became statistically non-significant in Model 3 (β=-0.080, 95% CI −0.164, 0.004; Fig. 2). In other pairwise comparisons, the Plant-based pattern was associated with a slower pace of aging compared to the Western with infrequent fish pattern in Models 1 and 2 (β=-0.106, 95% CI −0.186, −0.026 and β=-0.085, 95% CI −0.164, −0.006, respectively; results not shown). The strength of this association continued to decrease and became statistically non-significant in Model 3 (β=-0.057, 95% CI −0.132, 0.018). Additionally, the pace of aging was slower in the Plant-based pattern compared to the Western with frequent fish pattern in Model 1 (β=-0.101, 95% CI −0.196, −0.006), but the effect sizes decreased in Models 2 and 3 (β=-0.059, 95% CI −0.153, 0.035 and β=-0.039, 95% CI −0.127, 0.050, respectively).

The associations between dietary patterns and biological aging stratified by sex are presented in Supplementary Figures (Fig. S2, Fig. S3). Regarding AAPC-Grim, no major differences were observed between the sexes, and the effect sizes were approximately similar to those observed in the whole sample. However, when DunedinPACE was used as an outcome variable, the associations were stronger among males than females. Among males, the pace of aging was significantly slower in the Balanced-average pattern compared to the High fast food, low F&V pattern in all Models (β=-0.164, 95% CI −0.283, −0.045; β=-0.121, 95% CI −0.235, −0.007; and β=-0.123, 95% CI −0.232, −0.013 in Models 1, 2, and 3, respectively). In contrast, most effect sizes among females were close to zero.

We also tested for potential interactions in the individual-level analyses in Model 3 to evaluate whether sex- or lifestyle-related covariates may modify the association between diet and biological aging. An interaction was observed between BMI and dietary patterns in a model with AAPC-Grim (p<0.001; Fig. S4). Since it was thought that the association between BMI and biological aging could be curvilinear, we conducted a sensitivity analysis in which the quadratic term of BMI and its interactions with diet were included in the model. An interaction between the quadratic term and dietary patterns was observed (p=0.006), indicating a curvilinear association that varies across dietary patterns (Fig. 3). At low levels of BMI, biological aging was accelerated in High fast food, low F&V pattern compared to other dietary patterns. The interaction term between BMI and diet was non-significant in a model with DunedinPACE (p=0.350). The association between diet and both of the epigenetic aging measures was independent of sex, nonalcoholic energy intake, smoking status, alcohol consumption, and the Baecke sport index (p for interaction>0.05 in all).

**Fig. 3.**
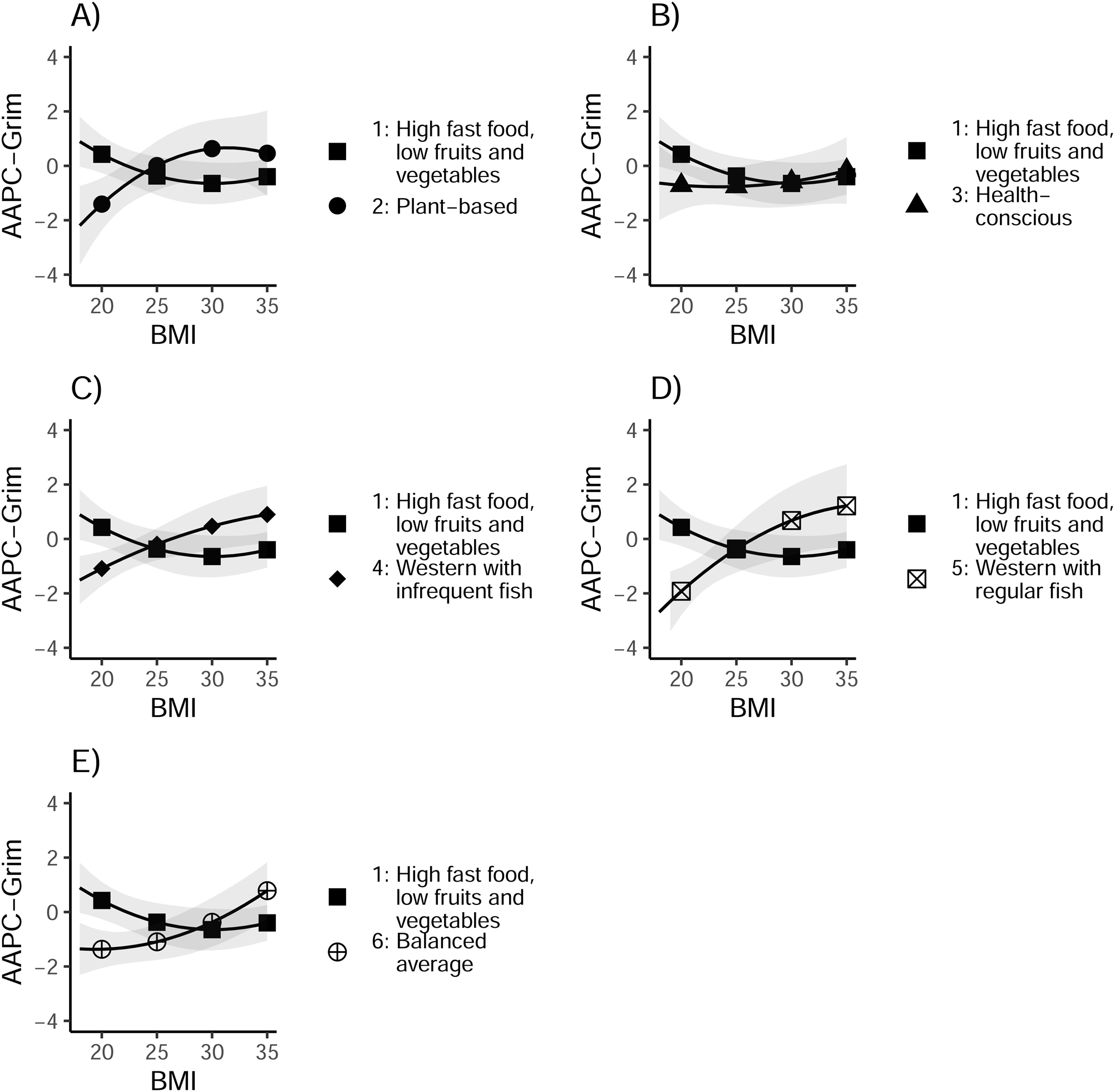
Differences in principal component-based GrimAge acceleration at different BMI levels between dietary patterns in comparison to the High fast food, low fruits and vegetables (A–E). The estimated means were obtained from the linear regression model, and the gray areas depict the 95% confidence intervals. AAPC-Grim, principal component-based GrimAge acceleration; BMI, body mass index.

Furthermore, we examined the similarity of the MZ and DZ twin pairs in terms of dietary pattern membership by comparing the proportions of concordant and discordant twin pairs. Among the MZ pairs, the ratio of the proportions of concordant twin pairs for dietary pattern membership to discordant pairs was 0.78 (43.9%/56.1%). The corresponding ratio among the DZ pairs was 0.32 (24.3%/75.7%). The higher proportion of concordant pairs among the MZ compared to the DZ twin pairs suggests genetic influences on dietary habits. The results of the within-pair analyses are presented in Figs. 4 and 5. The associations observed in the individual-level analyses were replicated in the within–twin pair analyses conducted for all twin pairs and DZ twin pairs; however, the effect sizes tended to be smaller for the MZ twin pairs, particularly in our fully adjusted models.

**Fig. 4.**
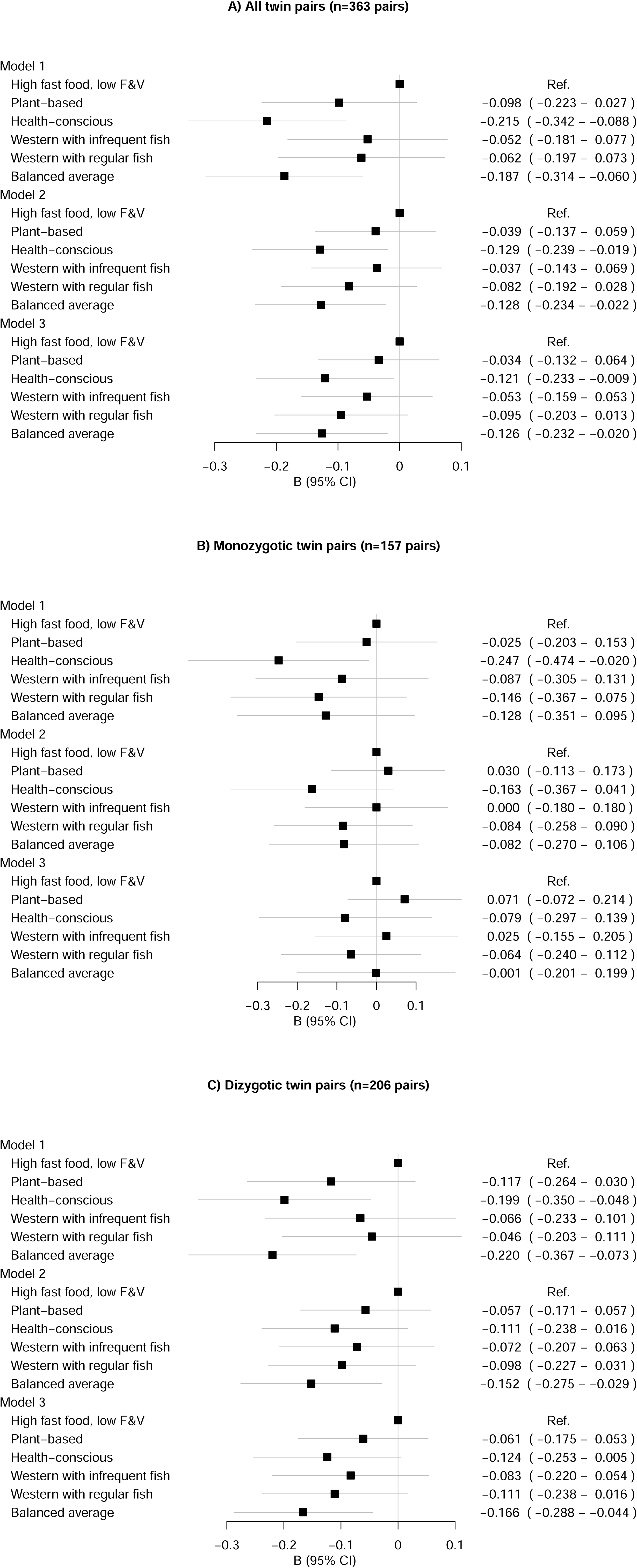
Differences in principal component-based GrimAge acceleration between dietary patterns in A) all pairs, B) monozygotic twin pairs, and C) dizygotic twin pairs: within-pair analysis. Model 1 was adjusted for family relatedness, total nonalcoholic energy intake, and sex; Model 2 was adjusted for the covariates in Model 1 plus smoking status and daily alcohol intake; and Model 3 was adjusted for the covariates in Model 2 plus BMI and the Baecke sport index. F&V, fruits and vegetables; B, standardized regression coefficient; CI, confidence interval; Ref., reference group.

**Fig. 5.**
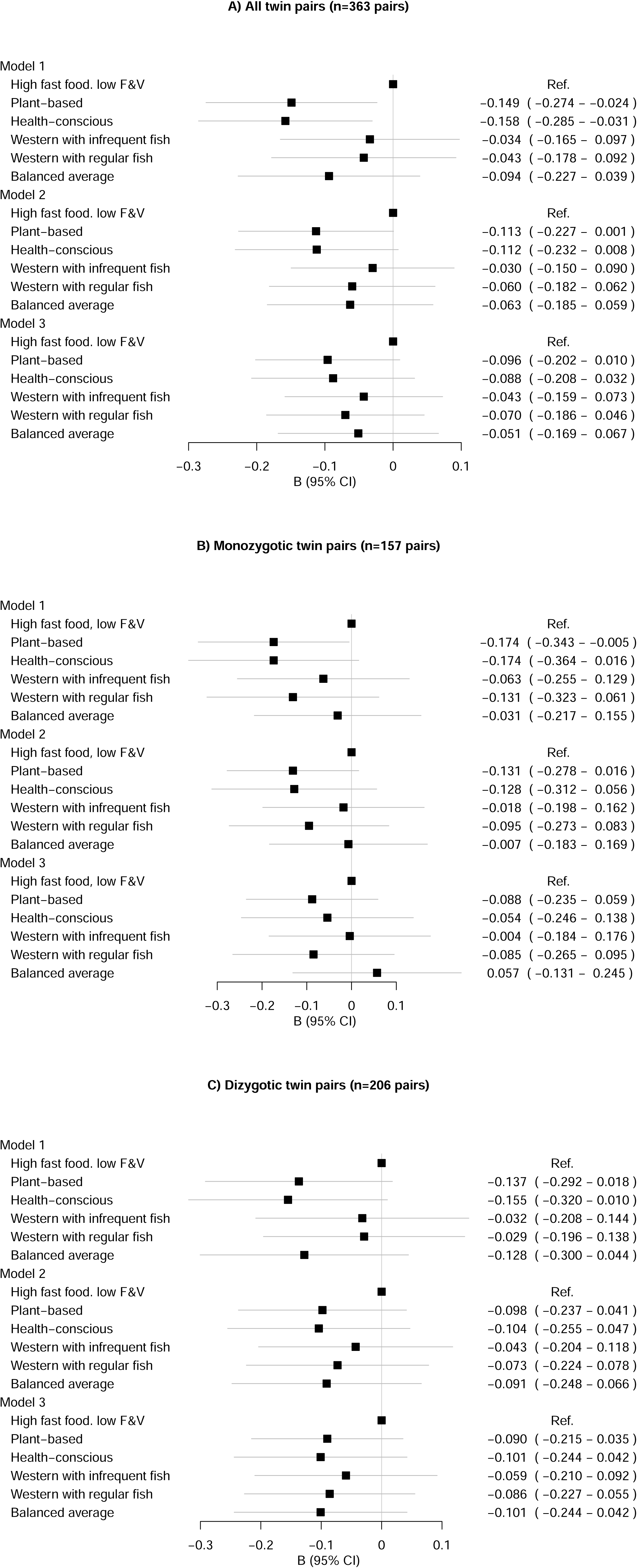
Differences in the pace of aging (measured by the DunedinPACE estimator) between dietary patterns in A) all pairs, B) monozygotic twin pairs, and C) dizygotic twin pairs: within-pair analysis. Model 1 was adjusted for family relatedness, total nonalcoholic energy intake, and sex; Model 2 was adjusted for the covariates in Model 1 plus smoking status and daily alcohol intake; and Model 3 was adjusted for the covariates in Model 2 plus BMI and the Baecke sport index. F&V, fruits and vegetables; B, standardized regression coefficient; CI, confidence interval; Ref., reference group.

## DISCUSSION

In this study, we investigated the associations between LCA-derived dietary patterns and biological aging among young adult twin pairs, both at the individual and within–twin pair levels. In general, diets emphasizing higher consumption of fruits and vegetables and lower intakes of meat, fast food, and sugar-sweetened beverages were associated with slower biological aging, while diets low in fruits and vegetables and high in meat, fast food, and sugar-sweetened beverages were linked to faster biological aging. However, the strengths of the associations were significantly attenuated when additional lifestyle factors were considered covariates. The associations between diets and biological aging differed according to sex when DunedinPACE was used to estimate biological aging but not when PC-GrimAge was applied. The associations found at the individual level were replicated at the within-pair level in all pairs and in the DZ pairs but were attenuated in the MZ pairs, suggesting the presence of genetic confounding.

Previous studies have mostly assessed diet quality using different dietary indices and have shown that higher-quality dietary patterns are associated with decreased biological aging (25–29,49,50). For example, two studies (25,26) found an association between lower Dietary Approaches to Stop Hypertension (DASH) scores (i.e., lower-quality diets) and age acceleration. Kresovich et al. (25) examined the links between the Healthy Eating Index, Alternative Healthy Eating Index, and Alternative Mediterranean diet with age acceleration and found inverse associations between all these metrics and age acceleration, indicating that higher diet quality is associated with slower age acceleration. In addition, Li et al. (27) found a link between higher scores of the Alternative Healthy Eating Index, the Mediterranean Dietary Score, and the Dietary Inflammatory Index with slower biological aging, both cross-sectionally and longitudinally. While the above-mentioned studies were conducted in middle-aged and older adults, Thomas et al. (28) reported that an association between a higher Mediterranean diet score and younger biological age was also observed among 20–40-year-old participants.

Dietary indices are, however, different from the LCA-derived dietary patterns used in this study. The former is a theory-based method, whereas LCA is a data-driven approach, therefore able to identify unobserved subgroups within a population instead of theory-driven index scores. Thus, our results are not directly comparable to those of earlier studies. However, the dietary patterns associated with slower biological aging in our study have similar characteristics to those with high dietary index scores, such as high intake of vegetables and fruits and low intake of processed meat and sugar-sweetened beverages.

One potential mechanism that may underlie the association between a healthier diet and slower biological aging could involve the intake of polyphenols. Polyphenols (i.e., phenolic acids, flavonoids, stilbenoids, and lignans) are found mainly in vegetables, fruits, berries, nuts, herbs, soy, tea, coffee, cocoa, and olive oil (51). Polyphenols have been reported to cause both direct and indirect modifications to the levels and activity of DNA methyltransferases (DNMTs) (52). DNMTs are enzymes that transfer a methyl group to the 5-carbon position of cytosine and are thus responsible for both de novo methylation processes and their maintenance (53). Hence, diets rich in polyphenols may result in favorable changes in DNAm levels and at least partly explain the slower age acceleration observed in patterns with high intakes of vegetables and fruits.

Moreover, high consumption of red meat is associated with accelerated biological aging when estimated by the Horvath clock, PhenoAge, and GrimAge metrics (20,54,55). Consistent with these findings, we also observed that dietary patterns rich in red meat and processed red meat (e.g., salami, sausage dishes) were associated with faster biological aging compared to diets with lower meat intake. Potential mechanisms behind these associations include the detrimental effects of high intakes of heme iron and the presence of heterocyclic amines (HAAs) and polycyclic aromatic hydrocarbons (PAHs) found on the surface of charred meat, all of which have been suggested to have carcinogenic effects (56). Additionally, saturated fats found in meat, as well as sodium and nitrates found in processed red meat, have been associated with an increased risk of cardiovascular diseases and cancer (56,57). However, it was not within the scope of our study to investigate the role of a single food item in biological aging. In fact, since high meat consumption coincided with low vegetable intake and high consumption of sugar-sweetened beverages, it is impossible to isolate the specific role of red meat consumption from other dietary characteristics.

It has been shown that modifiable lifestyle habits, such as smoking, alcohol consumption, physical activity, and dietary choices, tend to cluster among both adolescents and adults (58–61). In line with this finding, we found that smoking status, alcohol intake, BMI, and sports participation varied across the different dietary patterns. Therefore, as seen in Figs. 2, 4, and 5, the strengths of the observed associations between diet and biological aging decreased after adjusting for other lifestyle factors. This suggests that although dietary factors may be at least partially independently associated with biological aging, some associations appear to be confounded by other aspects of lifestyle. For example, Thomas et al. (28) reported that the associations between a healthy diet and physical activity with younger biological age were additive, in that low diet quality can be partially compensated by higher physical activity levels. In addition, obesity was associated with older biological age compared to normal weight, even in participants with a high diet quality (28).

Regarding sex differences, we found that the associations between diet and biological aging did not differ between males and females when PC-GrimAge was used as an outcome variable. However, we found a difference between sexes regarding the pace of aging, with the association being strong in males and nonexistent in females after adjusting for other lifestyle factors. This finding is in contrast to the results of a previous study (28), which found that the effect sizes in the association between the Mediterranean Diet score and the PhenoAge metric were slightly stronger in females than in males. As few studies have investigated the correlation between diet and biological aging separately for both sexes, further studies with larger sample sizes and different biological aging metrics should address this issue.

Interestingly, we found an interaction between BMI and dietary patterns in the AAPC-Grim model. More specifically, among individuals with low BMI values, biological aging was accelerated in the High fast food, low F&V group compared to all other groups, whereas among those with higher BMI values (especially those with overweight or obesity), AAPC-Grim yielded lower values in the High fast food, low F&V group. Our results suggest that among individuals with diets characterized by low consumption of vegetables, fruits, and berries and relatively high intake of meat, fast foods, and sugar-sweetened beverages, a lower BMI may be associated with greater age acceleration compared to individuals with similar diets but higher BMIs. The mechanisms behind this phenomenon remain elusive but may be related to low or inadequate intake of essential nutrients in those with low BMI. At the same time, we cannot exclude the possibility that some confounders were not addressed in our analysis. For instance, genetic predispositions, certain diseases, stress, socioeconomic status, or unmeasured lifestyle factors may have influenced our findings. However, the results indicate a synergistic influence of diet and BMI on biological aging, at least when AAPC-Grim is used as the metric for biological aging.

Our findings of more similar dietary patterns among MZ twins compared to DZ twins align with the results of earlier studies showing that among adults, genetic factors explain dietary patterns more than the shared childhood and youth environment (62–64). In addition, our finding that the associations observed in the individual-level analyses were replicated at the pairwise level among all pairs and among the DZ pairs suggests that shared environmental factors might not explain the observed associations between diet and biological aging. In contrast, the smaller effect sizes in the MZ twin pairs compared with the DZ twin pairs suggest that the relationship between diet and biological aging may be at least partially confounded by genetics.

Our study is not without limitations. First, the data were collected at one time point, thus limiting the interpretation of causal associations. However, reverse causality seems unlikely to explain our findings, as biological aging is not expected to influence dietary habits. Instead, residual confounding likely influenced our results, as demonstrated by the decreased effect sizes with the addition of covariates. The possibility that some residual confounders may still affect our fully adjusted models cannot be excluded. Second, the study participants were all of European ancestry based on genetic testing, which may limit the generalizability of our results to other ethnic groups. Third, dietary intakes were self-reported, which raises the possibility of misreporting. Finally, our study had a relatively small sample size, particularly in the Plant-based pattern, which may have led to limited statistical power to detect differences between dietary patterns. However, the strengths of our study include a high response rate, the utilization of a population-based cohort group, a twin study design, the use of the most recent biological aging metrics, and a person-centered approach to analyzing dietary patterns (i.e., LCA).

## CONCLUSION

Our results suggest that diets low in fruits and vegetables and high in meat, fast food, and soft drinks are already associated with faster biological aging in young adulthood compared with diets high in fruits and vegetables and low in processed red meat and sugar-sweetened beverages. The findings of this study indicate that adhering to the recommended dietary habits from a young age may mitigate age-related health risks later in life. However, sex differences, variations in the metrics used to measure biological aging, clustering effects of healthy and unhealthy lifestyle habits on biological aging, and genetic confounding need to be considered when interpreting the findings.

## Acknowledgments

We acknowledge all the participants involved in this study.

## Author contributions

SR and ES designed the study. SR and AK designed and conducted the statistical analysis and interpreted and visualized the data. JK, KHP, and MO designed and collected the FinnTwin12 data. LB prepared the dietary data. AH preprocessed the DNAm data and computed the biological aging estimates. SR drafted the first version of the manuscript, and AK and ES made important contributions through their edits and suggestions. All authors read, revised, and approved the final manuscript.

## Data statement

Data are available for qualified researchers through a standardized application procedure (see the website (https://thl.fi/en/research-and-development/thl-biobank/for-researchers) for details.

---

Fig. S1. Probabilities of consumption for food items treated as binary variables by dietary patterns derived from latent class analysis.

Fig. S2. Differences in principal component-based GrimAge acceleration between dietary patterns among A) males and B) females: individual-level analysis.

Model 1 was adjusted for family relatedness, total nonalcoholic energy intake, and sex; Model 2 was adjusted for the covariates in Model 1 plus smoking status and daily alcohol intake; and Model 3 was adjusted for the covariates in Model 2 plus BMI and the Baecke sport index.

F&V, fruits and vegetables; B, standardized regression coefficient; CI, confidence interval; Ref., reference group.

Fig. S3. Differences in the pace of aging (measured by the DunedinPACE estimator) between dietary patterns among A) males and B) females: individual-level analysis.

Model 1 was adjusted for family relatedness, total nonalcoholic energy intake, and sex; Model 2 was adjusted for the covariates in Model 1 plus smoking status and daily alcohol intake; and Model 3 was adjusted for the covariates in Model 2 plus BMI and the Baecke sport index.

F&V, fruits and vegetables; B, standardized regression coefficient; CI, confidence interval; Ref., reference group.

Fig. S4. Differences in principal component-based GrimAge acceleration at different BMI levels between dietary patterns in comparison to the High fast food, low fruits and vegetables pattern (A– E). The estimated means were obtained from the regression model with quadratic terms of BMI, and the gray areas depict the 95% confidence intervals.

AAPC-Grim, principal component-based GrimAge acceleration; BMI, body mass index.

