## Supplementary material for "Suboptimal dietary patterns are associated with accelerated biological aging in young adulthood: a twin study": Table S1

Table S1. Model fit of the latent class models with 1–7 classes based on dietary intake data (n=826).

| Classes | AIC | BIC | aBIC | VLMR-LRT | LMR | Entropy | Class sizes | AvePP |
| --- | --- | --- | --- | --- | --- | --- | --- | --- |
| 1 | 120564 | 121045 | 120721 | - | - | - | - |  |
| 2 | 117059 | 118026 | 117375 | <0.001 | <0.001 | 0.911 | 53.1%, 46.9% | 0.98, 0.98 |
| 3 | 115140 | 116592 | 115614 | 0.003 | 0.003 | 0.917 | 40.4%, 30.3%, 29.4% | 0.97, 0.97, 0.96 |
| 4 | 114079 | 116018 | 114713 | 0.273 | 0.274 | 0.925 | 29.9%, 29.5%, 27.8%, 12.7% | 0.97, 0.96, 0.96, 0.97 |
| 5 | 113411 | 115835 | 114203 | 0.770 | 0.770 | 0.927 | 27.2%, 26.3%, 25.5%, 12.5%, 8.5% | 0.95, 0.95, 0.97, 0.96, 0.98 |
| 6 | 112875 | 115785 | 113826 | 0.802 | 0.802 | 0.934 | 23.5%, 20.2%, 19.9%, 14.4%, 13.4%, 8.6% | 0.95, 0.96, 0.95, 0.96, 0.98, 0.99 |
| 7 | 112870 | 116266 | 113979 | 0.760 | 0.760 | 0.928 | 20.9%, 16.2%, 14.5%, 14.1%, 13.1%, 12.8%, 8.3% | 0.93, 0.94, 0.97, 0.97, 0.93, 0.95, 0.97 |
| AIC, Akaike’s information criterion; BIC, Bayesian information criterion; aBIC, sample size-adjusted Bayesian information criterion; VLMR, Vuong–Lo–Mendell–Rubin likelihood ratio test; LMR-LRT, Lo–Mendell–Rubin-adjusted likelihood ratio test; AvePP, average posterior probabilities for most likely latent class membership. | | | | | | | | |
