## Supplementary material for "Suboptimal dietary patterns are associated with accelerated biological aging in young adulthood: a twin study": Table S2

Table S2. Demographic and dietary characteristics of participants with different dietary patterns.

|  | Pattern 1 | Pattern 2 | Pattern 3 | Pattern 4 | Pattern 5 | Pattern 6 |
| --- | --- | --- | --- | --- | --- | --- |
|  | High fast food, low F&V (n=116) | Plant-based (n=71) | Health-conscious (n=168) | Western with infrequent fish (n=194) | Western with regular fish (n=111) | Balanced average (n=166) |
| Age, years, mean (SD) | 22.5 (0.7) | 22.3 (0.7) | 22.4 (0.6) | 22.5 (0.7) | 22.4 (0.7) | 22.3 (0.7) |
| Sex |  |  |  |  |  |  |
| Males, n (%) | 57 (49.1) | 7 (9.9) | 34 (20.2) | 116 (59.8) | 97 (87.4) | 34 (20.5) |
| Females, n (%) | 59 (50.9) | 64 (90.1) | 134 (79.8) | 78 (40.2) | 14 (12.6) | 132 (79.5) |
| BMI, kg/m^2^, median (25th, 75th percentile) | 22.4 (21.0, 25.3) | 21.9 (20.3, 24.2) | 22.6 (20.9, 25.1) | 23.1 (21.1, 25.6) | 23.3 (21.2, 25.2) | 22.3 (20.4, 24.3) |
| The Baecke sport index, mean (SD) | 2.6 (0.8)^a^ | 3.1 (0.7)^b^ | 3.3 (0.8)^c^ | 2.8 (0.7)^d^ | 3.1 (0.7)^e^ | 3.0 (0.7)^f^ |
| Smoking status |  |  |  |  |  |  |
| Never, n (%) | 56 (48.3) | 46 (64.8) | 105 (62.5) | 103 (53.1) | 34 (30.6) | 94 (56.6) |
| Former, n (%) | 7 (6.0) | 4 (5.6) | 18 (10.7) | 11 (5.7) | 14 (12.6) | 15 (9.0) |
| Occasionally, n (%) | 15 (12.9) | 8 (11.3) | 15 (8.9) | 19 (9.8) | 14 (12.6) | 20 (12.0) |
| Daily, n (%) | 38 (32.8) | 13 (18.3) | 30 (17.9) | 61 (31.3) | 49 (44.1) | 37 (22.3) |
| Alcohol intake, g/d, median (25th, 75th percentile) | 9.9 (3.0, 18.9) | 8.3 (4.6, 11.6) | 8.3 (3.3, 14.3) | 14.2 (6.6, 21.3) | 17.4 (8.4, 24.8) | 8.7 (4.6, 14.3) |
| Energy intake, kcal/d, mean (SD)^g^ | 1879 (626) | 1690 (467) | 1717 (366) | 2327 (595) | 2788 (543) | 2069 (410) |
| Carbohydrate intake, g/d, mean (SD) | 254 (92) | 244 (65) | 243 (59) | 302 (85) | 348 (69) | 283 (68) |
| Carbohydrate intake, E%, mean (SD) | 49.3 (7.2) | 53.2 (6.2) | 51.4 (5.3) | 47.5 (4.7) | 45.9 (4.0) | 50.3 (4.5) |
| Protein intake, g/d, mean (SD) | 76 (29) | 66 (23) | 75 (18) | 97 (27) | 126 (29) | 86 (19) |
| Protein intake, E%, mean (SD) | 15.5 (3.2) | 15.0 (2.9) | 16.9 (2.3) | 15.9 (2.2) | 17.2 (2.0) | 16.1 (2.2) |
| Fat intake, g/d, mean (SD) | 66 (27) | 56 (21) | 55 (15) | 86 (24) | 105 (26) | 71 (16) |
| Fat intake, E%, mean (SD) | 30.2 (5.9) | 28.3 (5.2) | 27.8 (4.7) | 31.9 (4.2) | 32.1 (3.6) | 30.1 (4.1) |

^a^ n=114; ^b^ n=67; ^c^ n=157; ^d^ n=193; ^e^ n=108; ^f^ n=161; ^g^ from non-alcoholic foods and beverages

F&V, fruits and vegetables; SD, standard deviation; BMI, body mass index; E%, energy percentage.
