## Supplementary figures and images for "Suboptimal dietary patterns are associated with accelerated biological aging in young adulthood: a twin study"

### Fig. S1

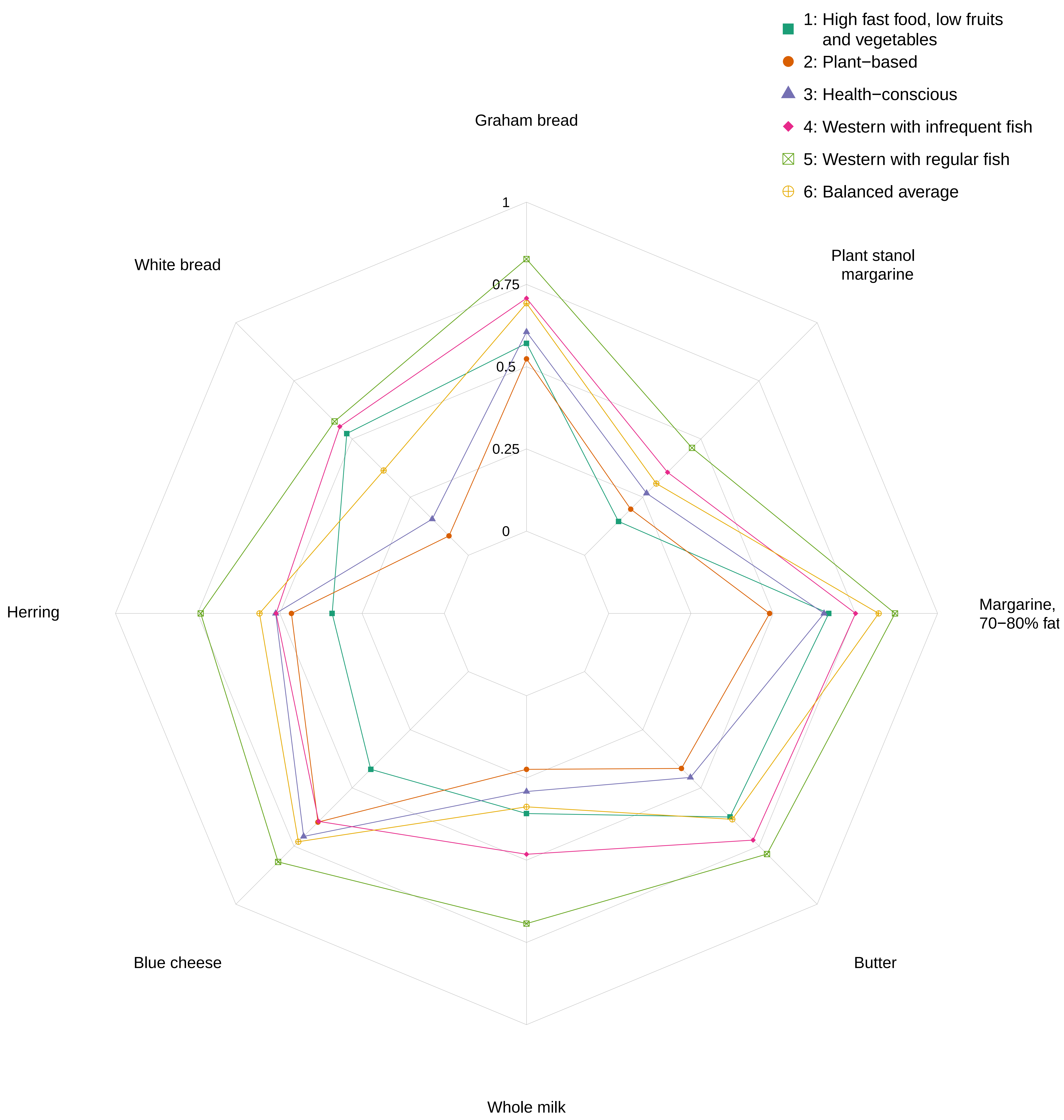

### Fig. S2

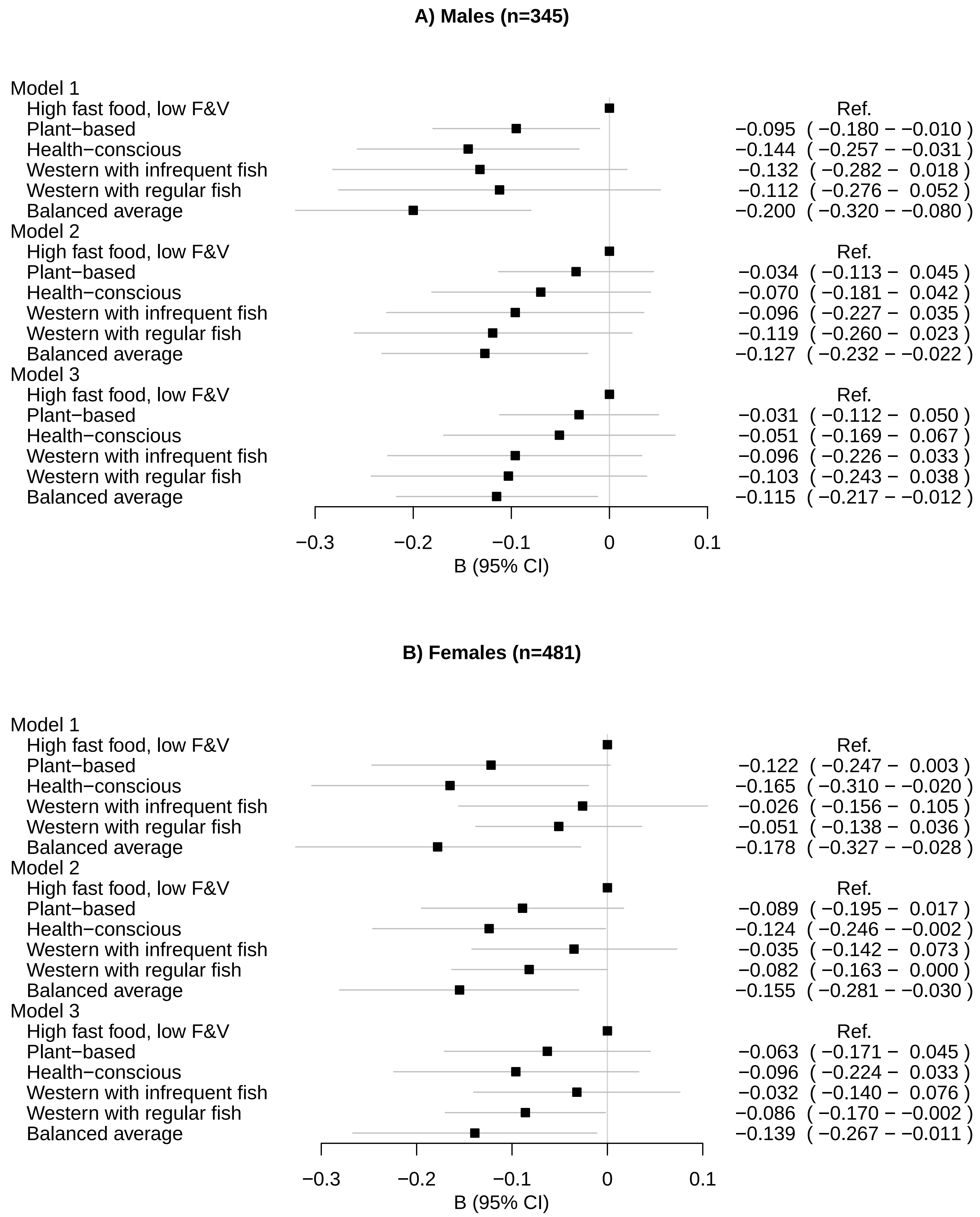

### Fig. S3

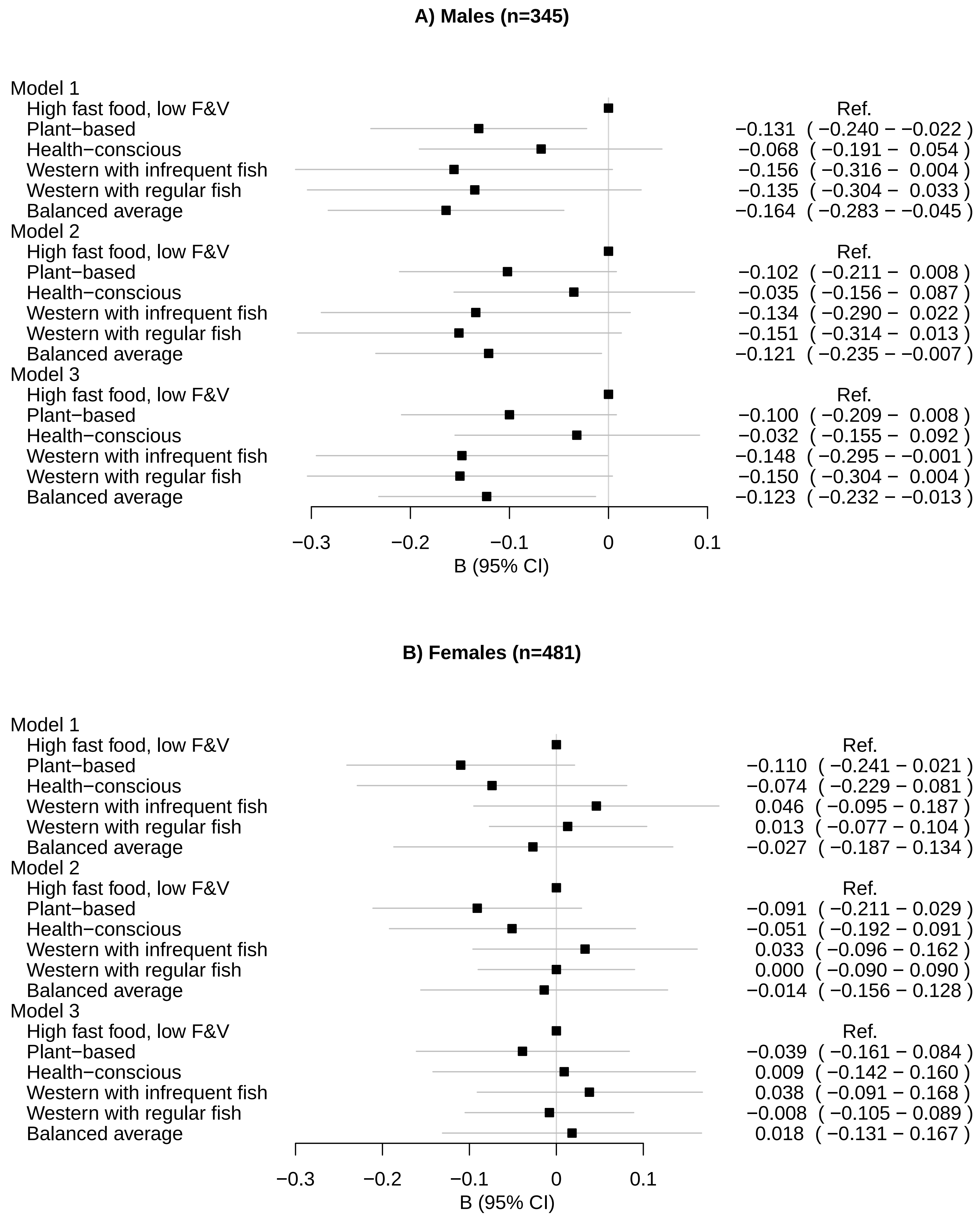

### Fig. S4

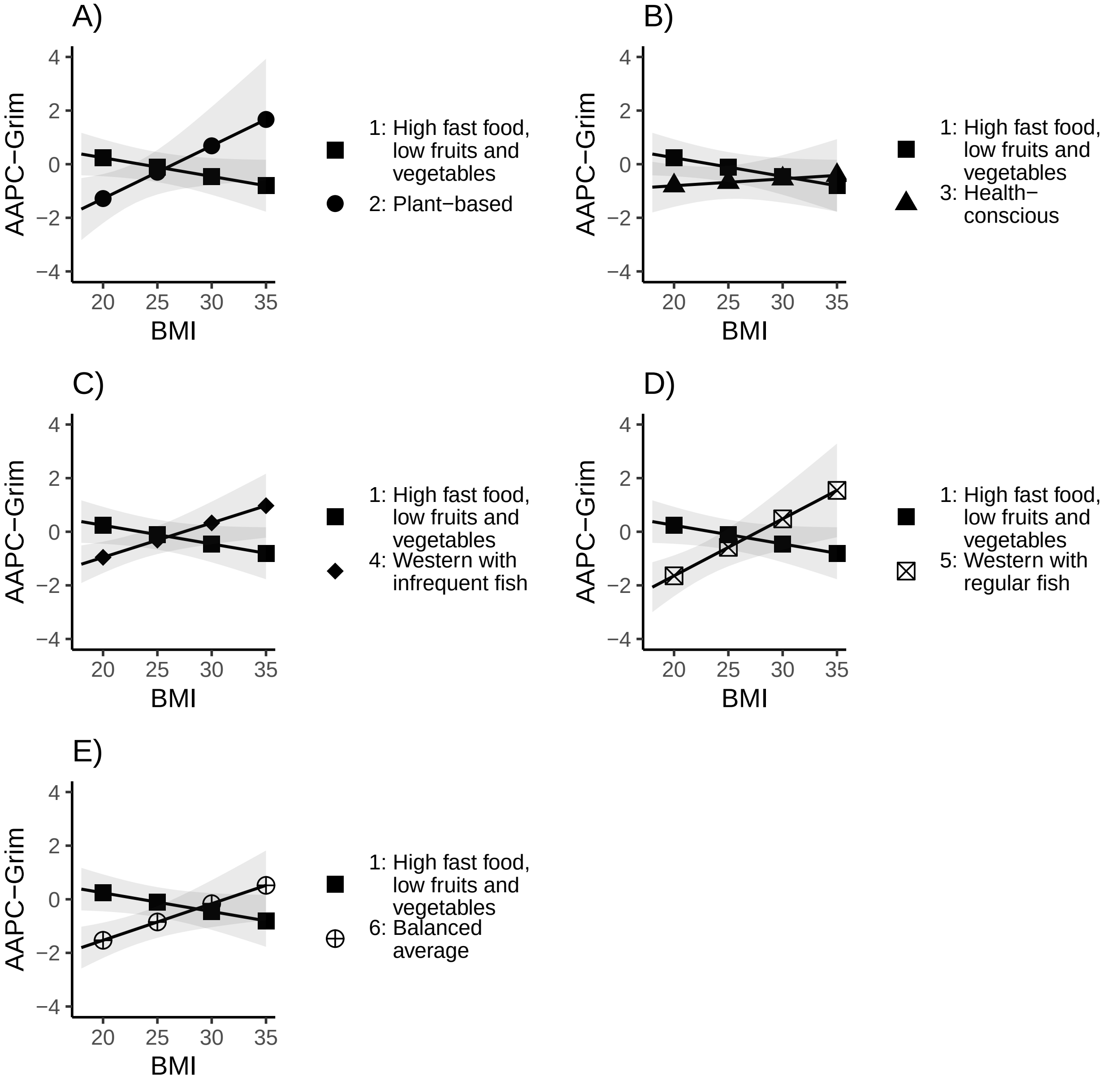
